# COVID-19 vaccination coverage by company size and the effects of socioeconomic factors and workplace vaccination in Japan: a cohort study

**DOI:** 10.1101/2022.03.30.22273203

**Authors:** Koji Mori, Takahiro Mori, Tomohisa Nagata, Hajime Ando, Ayako Hino, Seiichiro Tateishi, Mayumi Tsuji, Keiji Muramatsu, Yoshihisa Fujino, the CORoNa Work Project

## Abstract

**Background:** Vaccination is considered the most effective control measure against COVID-19. Vaccine hesitancy and equitable vaccine allocation are important challenges to disseminating developed vaccines. To promote COVID-19 vaccination coverage, the government of Japan established the workplace vaccination program. However, while it appears that the program was effective in overcoming vaccine hesitancy, the program may have hindered the equitable allocation of vaccines because it mainly focused on employees of large companies. We investigated the relationship between company size and COVID-19 vaccination completion status of employees and the impact of the workplace vaccination program on this relationship.

**Methods:** We conducted an internet-based prospective cohort study from December 2020 (baseline) to December 2021. The data were collected using a self-administered questionnaire survey. Briefly, 27,036 workers completed the questionnaire at baseline and 18,560 at follow-up. After excluding ineligible respondents, we finally analyzed the data from 15,829 participants. At baseline, the participants were asked about the size of the company they worked for, and at follow-up they were asked about the month in which they received their second COVID-19 vaccine dose and the availability of a company-arranged vaccination opportunity.

**Results:** In each month throughout the observation period, the odds of having received a second COVID-19 vaccine dose were significantly lower for small-company employees than for large-company employees in the sex- and age-adjusted model. This difference decreased after adjusting for socioeconomic factors, and there was no significant difference after adjusting for the availability of a company-arranged vaccination opportunity.

**Conclusions:** The workplace vaccination program implemented in Japan to control the COVID-19 pandemic may have been effective in overcoming vaccine hesitancy in workers; however, it may have caused an inequitable allocation of vaccines between companies of different sizes. Because people who worked for small companies were less likely to be vaccinated, it will be necessary to enhance support of vaccination for this population in the event of future infectious disease outbreaks.

**Trial registration:** Not applicable.

## Introduction

Vaccination programs are underway worldwide because vaccination is the most effective measure to control the coronavirus disease 2019 (COVID-19) pandemic, which was declared by the World Health Organization (WHO) in March 2020. Since the outbreak of COVID-19 in China in December 2019 [1], various types of vaccines have been developed in a short period of time [2]. Some of these are mRNA vaccines, representing a new type of vaccine technology [3].

Disseminating vaccines presents many challenges, among which vaccine hesitancy and equitable allocation are prominent. Vaccine hesitancy, defined as the “delay in acceptance or refusal of vaccination despite the availability of vaccination services” is considered a major public health challenge in infectious disease control because it delays vaccination of the population and inhibits the acquisition of herd immunity [4]. Various factors, including socioeconomic [5] and psychological factors [4], have been found to contribute to people’s vaccine hesitancy. Such factors have also been examined in the context of COVID-19 vaccination [6.7]. The equitable allocation of vaccines is based on maintaining equity in the order of vaccination according to risk regardless of social status, for example by starting with healthcare workers and those at higher risk of serious illness [8].

In Japan, the majority of the population had some level of initial vaccine hesitancy to receive a COVID-19 vaccine [9,10]. Nevertheless, by the end of December 2021, approximately 80% of the population had received two vaccine doses [11]. In Japan, COVID-19 vaccination efforts began on February 17, 2021 using two mRNA vaccines: one from Pfizer Inc. and one from Moderna Inc. In consideration of equitable vaccine allocation, the vaccination of healthcare workers was followed by the vaccination of older adults [12]. Thereafter, vaccination progressed through the general population in stages according to age.

An aspect of COVID-19 vaccination in Japan has been the availability of vaccination at workplaces in addition to community settings provided by municipalities and clinics [12]. Compared with other developed countries, the start of the vaccination program was delayed in Japan. To make up for this delay, the government appointed a minister to be in charge and set a goal of administering one million vaccinations per day. Part of the vaccination strategy was to implement the opportunity for workplace vaccination, which was conducted mainly by occupational health professionals such as occupational physicians and occupational health nurses. As a result, 9,654,000 people received their second vaccine dose through the workplace vaccination program, which started on June 21, 2021 [13]. Workplace vaccination, which provides a convenient vaccination opportunity, may have reduced vaccine hesitancy because several psychological and social factors can positively influence a person’s vaccination decision.

The workplace COVID-19 vaccination program in Japan, however, may have negatively affected the equitable allocation of vaccine doses. This program primarily targeted large companies, with a minimum of 2,000 doses to be delivered to a single location (i.e., an expected vaccination coverage of at least 1,000 persons [13]). Thus, there were barriers to its implementation in small and medium-sized companies. Therefore, company size may have affected the timing and coverage of employees receiving the second COVID-19 vaccine dose.

We hypothesized that while the workplace vaccination program facilitated COVID-19 vaccination, there was a size-dependent difference among companies in the timing of employees receiving the second vaccine dose and that this difference was influenced by the availability of a company-arranged vaccination opportunity. In a survey conducted in Japan during the COVID-19 pandemic, there were differences in the implementation of infection control measures and the opportunity to work remotely depending on the size of the company [14,15]. Disparities in occupational health measures, such as workplace environmental and health measures, have arisen and depend on the size of the company. Such disparities have also been found in the establishment of COVID-19 countermeasures. Therefore, rather than the government’s workplace vaccination program ensuring vaccine equity, this program may have increased disparities in infection risk because of differences in the completion of COVID-19 vaccination based on company size.

We conducted a prospective cohort study to examine the relationship between company size and COVID-19 vaccination completion and the impact of the workplace vaccination program on this relationship, focusing on the period between July and December 2021, when the general population in Japan was receiving the second vaccine dose.

## Methods

### Study design and participants

This study was a part of the Collaborative Online Research on Novel-coronavirus Work Study (the CORoNa Work Study) and was conducted using a prospective cohort study design. The survey was commissioned to the internet survey company Cross Marketing Inc. (Tokyo Japan), and the data were collected using a self-administered online questionnaire. All participants gave informed consent, and the study was approved by the ethics committee of the University of Occupational and Environmental Health, Japan (approval number: R2-079 and R3-006).

The baseline survey was conducted from December 22 to 25, 2020. The protocol for the baseline survey has been previously reported in detail [16]. The participants were aged 20–65 years and were employed at the time of the baseline survey (N=33,087). Participants were included using cluster sampling by sex, age, region, and occupation. A total of 27,036 participants were included after excluding ineligible individuals: those for which no data on company size was available, who worked in the medical or welfare sectors, or who were older than 65 at the time of the follow-up survey.

The follow-up survey was conducted from December 15 to 22, 2021, 1 year after baseline. A total of 18,560 participants responded to the survey. Among them, respondents were excluded if they were unemployed, over 65 years of age, or employed in the health or welfare sector and thus eligible for priority vaccination at the time of the follow-up survey. Finally, 15,829 participants were included in the analysis. Figure 1 shows the flow diagram for this study.

**Figure 1.**
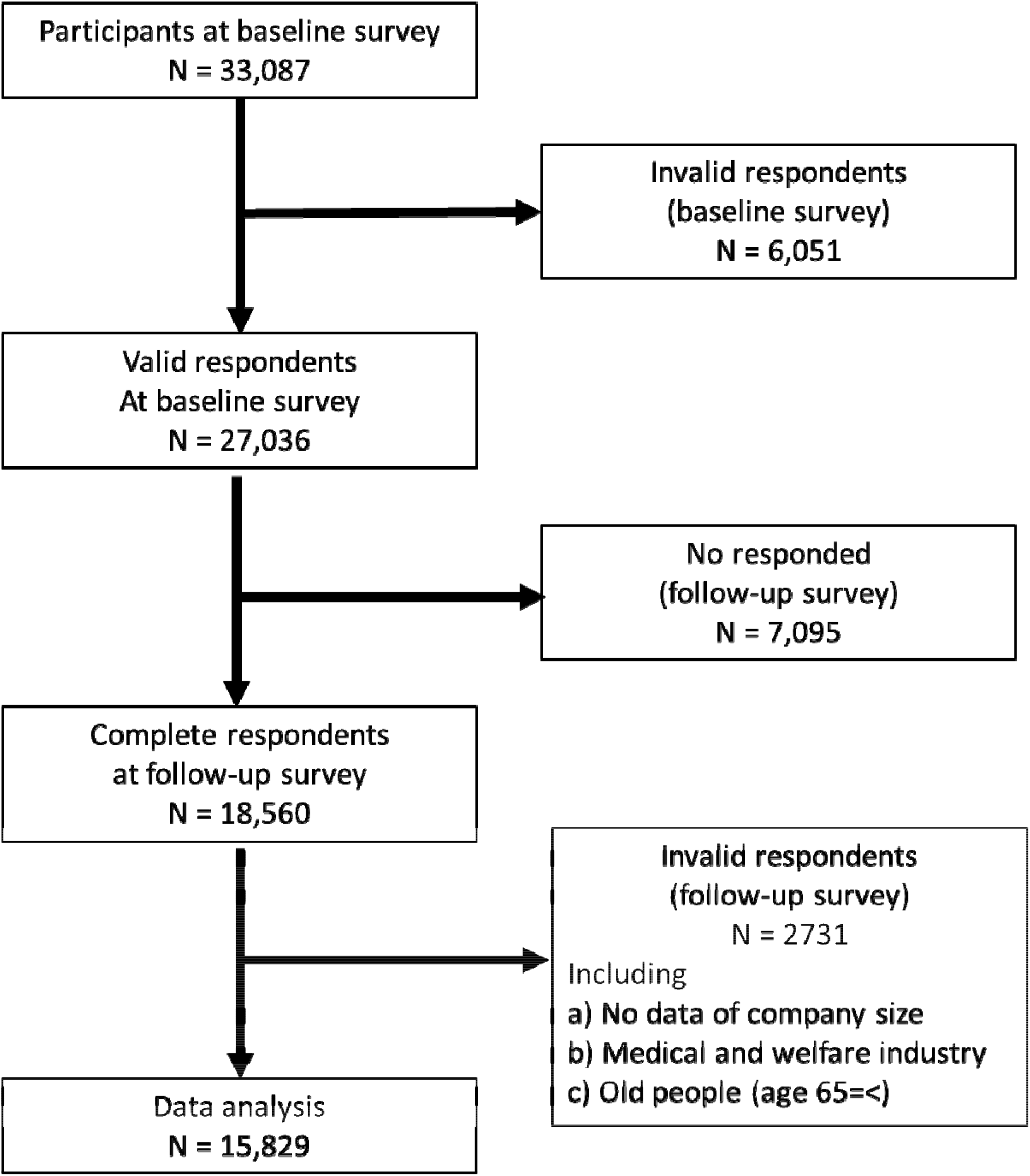
Flow diagram of the study participants.

### Second COVID-19 vaccination dose status

In the follow-up survey, we asked participants, “In what month did you receive the second COVID-19 vaccination?” Participants were requested to choose one of 12 options: the months of February 2021 through December 2021, or “have not received.” We then created a variable for completion status for each month after July. For example, completion by the end of September was defined as having received a second COVID-19 vaccine dose in any of the months from February through September. If a participant received the second vaccine dose in September, completion by July or August would be coded “no” but completion by September, October, November, and December would be coded “yes”.

### Company size

In the baseline survey, we asked participants, “How many employees are there at your company?” The participants could choose one of 10 options: 1 person (self-employed) or 2–4, 5–9, 10–29, 30–49, 50–99, 100–499, 500–999, 1000–9999, or 10,000 or more persons. We classified the responses into three categories: those who worked for small (1–49), medium-sized (50–999), or large (1,000 or more) companies. This classification was made because under the Industrial Safety and Health Act, the obligation to establish an occupational health management system differs depending on the size of the worksite [17]. Furthermore, the government-provided workplace vaccination program was eligible for locations that could vaccinate at least 1,000 people [13].

### Company-arranged vaccination opportunity

In the follow-up survey, we asked participants, “Has your company arranged an opportunity to receive the COVID-19 vaccine at the workplace, whether or not you took advantage of the opportunity?” Participants could choose one of three response options: yes, no, or unknown. We regarded “yes” to indicate that the vaccination opportunity was arranged, and the other answers to indicate that this was not arranged.

### Assessment of covariates

Participant characteristics were collected at baseline. The covariates included socioeconomic factors, occupation, and industry. Age was classified into five groups: 20–29, 30–39, 40–49, 50–59, and 60–65 years. Annual household income was classified into five categories: <2.00 million Japanese yen (JPY), 2.00–3.99 million JPY, 4.00–5.99 million JPY, 6.00–7.99 million JPY, and 8.00 million JPY or greater. Educational background was classified into three categories: junior high or high school, vocational school or college, and university or graduate school. Marital status was classified into three categories: married, divorced or widowed, and unmarried. Occupation was classified into 10 categories: general employee; manager; executive manager; public employee, faculty member, or non-profit organization employee; temporary or contract employee; self-employed; small office/home office; agriculture, forestry, or fishing; professional occupation (e.g., lawyer, tax accountant); and other occupations. Participants could choose one of 22 options for their work industry, which was then classified into nine categories based on the International Standard Industrial Classification of All Economic Activities: manufacturing, public service, information and communication, wholesale and retail, food service, education and religion, finance and insurance, construction, and others.

### Statistical analysis

The odds ratios (ORs) for the association between company size and completion of the second COVID-19 vaccine dose were estimated using a multilevel logistic model nested in the prefecture of residence to account for regional variability. The multivariate model was adjusted for sex and age (Model 1) and additionally adjusted for annual household income, educational background, marital status, occupation, and industry (Model 2). Finally, the model was adjusted for company-arranged vaccination opportunity (Model 3).

A *p*-value of less than 0.05 was considered statistically significant. All analyses were conducted using Stata (Stata Statistical Software: Release 16; StataCorp LLC, College Station, TX, USA).

## Results

Table 1 shows the participant characteristics by company size. Of the 15,829 participants, 4,272 (27%) worked for a large company, 5,117 (32%) for a medium-sized company, and 6,440 (41%) for a small company. As the company size increased, the percentage of participants with a high annual household income and a high educational background level increased. Furthermore, as the company size increased, the opportunity for company-arranged vaccination increased: 56% for large companies, 35% for medium-sized companies, and 14% for small companies.

**Table 1.** Participant characteristics according to company size

|  | Size of belonging company (Number of employees) |  |  |
| --- | --- | --- | --- |
|  | Large (1000 or more) | Medium (50-999) | Small (1-49) |
| Total | 4272 | 5117 | 6440 |
| Age |  |  |  |
| 20-29 | 185 (4.3%) | 259 (5.1%) | 173 (2.7%) |
| 30-39 | 611 (14.3%) | 764 (14.9%) | 769 (11.9%) |
| 40-49 | 1222 (28.6%) | 1601 (31.3%) | 1898 (29.5%) |
| 50-59 | 1755 (41.1%) | 1929 (35.7%) | 2511 (39.0%) |
| 60-65 | 499 (11.7%) | 664 (13.0%) | 1089 (16.9%) |
| Sex |  |  |  |
| Men | 2781 (65.1%) | 3172 (62.0%) | 3785 (58.8%) |
| Women | 1491 (34.9%) | 1945 (38.0%) | 2665 (41.2%) |
| Annual household income (million JPY) |  |  |  |
| <2 | 128 (3.0) | 226 (4.4%) | 660 (10.3%) |
| ≥2 and <4 | 514 (12.0%) | 990 (19.3%) | 1594 (24.8%) |
| ≥4 and <6 | 828 (19.4%) | 1316 (25.7%) | 1610 (25.0%) |
| ≥6 and <8 | 997 (23.3%) | 1068 (20.9%) | 1105 (17.2%) |
| ≥8 | 1805 (42.3%) | 1517 (29.6%) | 1471 (22.8%) |
| Educational background |  |  |  |
| Junior high or high school | 1018 (23.8%) | 1426 (27.9%) | 2117 (32.9%) |
| Vocational school, junior college or technical school | 613 (14.4%) | 1003 (19.6%) | 1575 (24.5%) |
| University or graduate school | 2641 (61.8%) | 2688 (52.5%) | 2748 (42.7%) |
| Marital status |  |  |  |
| Married | 1244 (29.1%) | 1704 (33.3%) | 2296 (35.7%) |
| Widowed/divorced | 339 (7.9%) | 432 (8.4%) | 755 (11.7%) |
| Never married | 2689 (62.9%) | 2981 (58.3%) | 3389 (58.3%) |
| Occupation |  |  |  |
| General employee | 2104 (49.3%) | 2773 (54.2%) | 2423 (37.6%) |
| Manager | 691 (16.2%) | 753 (14.7%) | 382 (5.9%) |
| Executive manager | 31 (0.7%) | 107 (2.1%) | 494 (7.7%) |
| Public employee, faculty member, or non-profit organization employee | 806 (18.9%) | 606 (11.8%) | 354 (5.5%) |
| Temporary/contract employee | 603 (14.1%) | 810 (15.8%) | 340 (5.3%) |
| Independent business (commercial and industrial services) | 9 (0.2%) | 15 (0.3%) | 1548 (24.0%) |
| Small office/home office | 3 (0.1%) | 3 (0.1%) | 270 (4.2%) |
| Agricultural, forestry, and fishing industries | 2 (0.0%) | 3 (0.1%) | 135 (2.1%) |
| Professional occupation (lawyer, tax accountant, etc.) | 11 (0.3%) | 14 (0.3%) | 140 (2.2%) |
| Other occupation | 12 (0.3%) | 33 (0.6%) | 354 (5.5%) |
| Industry |  |  |  |
| Manufacturing | 1069 (25.0%) | 1179 (23.0%) | 755 (11.7%) |
| Public service | 638 (14.9%) | 389 (7.6%) | 203 (3.2%) |
| Information and technology | 312 (7.3%) | 331 (6.5%) | 287 (4.5%) |
| Retail and wholesale | 244 (5.7%) | 355 (6.9%) | 629 (9.8%) |
| Eating/drinking | 110 (2.6%) | 182 (3.6%) | 513 (8.0%) |
| Education and religion | 275 (6.4%) | 417 (8.1%) | 507 (7.9%) |
| Finance | 434 (10.2%) | 213 (4.2%) | 161 (2.5%) |
| Construction | 96 (2.3%) | 139 (2.7%) | 428 (6.7%) |
| Other | 1064 (25.6%) | 1912 (37.4%) | 2957 (45.9%) |
| Vaccination arranged by company |  |  |  |
| Yes | 2383 (55.8%) | 1780 (34.8%) | 914 (14.2%) |
| No | 1889 (44.2%) | 3337 (65.2%) | 5526 (85.8%) |
| Month of 2nd COVID-19 vaccination |  |  |  |
| February-June | 161 (3.0%) | 205 (4.0%) | 197 (3.1%) |
| July | 450 (10.5%) | 467 (9.1%) | 607 (9.4%) |
| August | 1267 (29.7%) | 4354 (26.5%) | 1550 (24.1%) |
| September | 941 (22.0%) | 1140 (22.3%) | 1380 (21.4%) |
| October | 774 (18.1%) | 1063 (20.8%) | 1212 (18.8%) |
| November | 240 (5.6%) | 319 (6.2%) | 420 (6.5%) |
| December | 21 (0.5%) | 35 (0.7%) | 40 (0.6%) |
| Non-vaccinated | 418 (9.8%) | 534 (10.4%) | 1034 (16.1%) |

Table 2 shows the ORs for the association between company size and completion of the second COVID-19 dose by month. In the model adjusted only for age and sex (Model 1), participants who worked for a medium-sized company were significantly less likely to complete the second dose by August (OR=0.87, 95% CI: 0.79–0.94, p=0.001) and September (OR=0.86, 95% CI: 0.78–0.93, p<0.001) than those who worked for a large company. For small companies, the ORs decreased throughout the entire observation period, from July to December. In the model adjusted for the main socioeconomic factors (Model 2), the ORs for medium-sized and small companies tended to approach 1. For August and September, this tendency remained, but no significant difference was observed for the medium-sized companies. After adjusting for company-arranged vaccination opportunity (Model 3), the significant difference between small and large companies disappeared for the entire period analyzed. However, after October, participants who worked for medium-sized companies were significantly more likely to have received the second vaccine dose than those who worked for large companies (OR=1.14, 95% CI: 1.01–1.28, p=<0.029). In each month throughout the observation period, those who had a company-arranged vaccination opportunity were significantly more likely to have received the second vaccine dose.

**Table 2.** Association between company size and completion of the second COVID-19 vaccine dose

| Second vaccination |  | model 1 |  |  | model 2 |  |  | model 3 |  |
| --- | --- | --- | --- | --- | --- | --- | --- | --- | --- |
| Comp. size | OD | 95%CI | P value | OD | 95%CI | P value | OD | 95%CI | P value |
| by July |  |  |  |  |  |  |  |  |  |
| Large | Ref. |  |  | Ref. |  |  | Ref. |  |  |
| Medium | 0.93 | 0.82-1.05 | 0.229 | 0.98 | 0.86-1.10 | 0.691 | 1.06 | 0.94-1.21 | 0.330 |
| Small | 0.77 | 0.69-0.87 | <0.001 | 0.79 | 0.69-0.90 | 0.002 | 0.94 | 0.82-1.07 | 0.350 |
| Vaccination by comp. |  |  |  |  |  |  | 1.56 | 1.40-1.74 | <0.001 |
| by August |  |  |  |  |  |  |  |  |  |
| Large | Ref. |  |  | Ref. |  |  | Ref. |  |  |
| Medium | 0.87 | 0.79-0.94 | 0.001 | 0.92 | 0.84-1.00 | 0.065 | 1.05 | 0.96-1.15 | 0.277 |
| Small | 0.67 | 0.62-0.73 | <0.001 | 0.76 | 0.69-0.83 | <0.001 | 0.98 | 0.89-1.08 | 0.692 |
| Vaccination by comp. |  |  |  |  |  |  | 1.98 | 1.83-2.15 | <0.001 |
| by September |  |  |  |  |  |  |  |  |  |
| Large | Ref. |  |  | Ref. |  |  | Ref. |  |  |
| Medium | 0.86 | 0.78-0.93 | 0.001 | 0.92 | 0.84-1.01 | 0.085 | 1.04 | 0.95-1.15 | 0.349 |
| Small | 0.64 | 0.59-0.70 | <0.001 | 0.77 | 0.70-0.84 | <0.001 | 0.97 | 0.88-1.07 | 0.541 |
| Vaccination by comp. |  |  |  |  |  |  | 1.90 | 1.76-2.07 | <0.001 |
| by October |  |  |  |  |  |  |  |  |  |
| Large | Ref. |  |  | Ref. |  |  | Ref. |  |  |
| Medium | 0.92 | 0.82-1.02 | 0.127 | 1.00 | 0.89-1.12 | 0.971 | 1.14 | 1.01-1.28 | 0.029 |
| Small | 0.54 | 0.48-0.61 | <0.001 | 0.75 | 0.67-0.84 | <0.001 | 0.95 | 0.85-1.07 | 0.377 |
| Vaccination by comp. |  |  |  |  |  |  | 2.05 | 1.84-2.27 | <0.001 |
| by November |  |  |  |  |  |  |  |  |  |
| Large | Ref. |  |  | Ref. |  |  | Ref. |  |  |
| Medium | 0.93 | 0.81-1.10 | 0.218 | 1.01 | 0.88-1.15 | 0.918 | 1.15 | 1.01-1.32 | 0.042 |
| Small | 0.54 | 0.48-0.61 | <0.001 | 0.73 | 0.64-0.83 | <0.001 | 0.94 | 0.82-1.08 | 0.362 |
| Vaccination by comp. |  |  |  |  |  |  | 2.14 | 1.88-2.43 | <0.001 |
| by December |  |  |  |  |  |  |  |  |  |
| Large | Ref. |  |  | Ref. |  |  | Ref. |  |  |
| Medium | 0.94 | 0.82-1.07 | 0.329 | 1.02 | 0.89-1.17 | 0.759 | 1.17 | 1.02-1.35 | 0.029 |
| Small | 0.54 | 0.48-0.61 | <0.001 | 0.73 | 0.64-0.83 | <0.001 | 0.93 | 0.81-1.07 | 0.317 |
| Vaccination by comp. |  |  |  |  |  |  | 2.14 | 1.83-2.15 | <0.001 |

model 1: adjusted for age and sex
model 2: model 1+ adjusted for annual household income, education, marital status, occupation and industry
model 3: model 2 + adjusted for company-arranged vaccination
Size of company: large (1-49); medium-sized (50-999); large (1000 or more)
Vaccination by comp.: vaccination arranged by company

## Discussion

This study showed that employees of smaller companies were less likely to have received a second COVID-19 vaccine dose. In the months after the start of the workplace vaccination program, the second dose completion rate of participants who worked for medium-sized companies was lower than that of those who worked for large companies, but this difference disappeared later in the observation period. The significant difference in completion rate between small company employees and large company employees remained throughout the observation period. The difference between large and medium-sized company employees could mostly be explained by differences in socioeconomic factors. However, the difference between the small and large company employees could not be explained by those factors alone, although adjusting for the socioeconomic factors reduced the difference. After adjusting for company-arranged vaccination opportunity, the difference in employee second dose completion rate between large and small companies disappeared. Furthermore, medium-size companies had higher vaccine completion coverage than large companies in the latter half of the observation period.

The presence of vaccine hesitancy owing to a lack of trust in vaccination and other factors has been a challenge to achieving herd immunity through vaccination. Socioeconomic factors have been found to affect vaccination intention and uptake of other vaccines, such as the seasonal influenza vaccine [18] and the H1N1 vaccine [19]. The effects of socioeconomic factors on vaccination intention for the COVID-19 vaccine have also been examined [6,7]. Studies have generally found a positive association between vaccine uptake and annual income and educational background, although some studies have shown inverse associations [20, 21]. Several studies have found differences in willingness to vaccinate depending on one’s occupation and industry [22-24]. In the current study, after adjusting for socioeconomic factors, the difference in vaccination completion rate among employees of medium and large companies disappeared. After these adjustments, the difference between employees of small and large companies also became smaller. These findings suggest that socioeconomic factors affect the association between COVID-19 vaccination and company size in Japan.

In the present study, it was observed that participants who had a company-arranged vaccination opportunity were significantly more likely to have received the second vaccine dose, and after adjusting the model for the presence of a company-arranged vaccination opportunity, no significant difference in the second dose completion rate was found between employees of small and large companies for all months. These results suggests that the government’s implementation of the workplace vaccination program had a positive impact on the vaccination acceptance of employees who worked for companies that participated in the program. The company-arranged vaccination opportunities may have decreased vaccine hesitancy and increased vaccination coverage. To evaluate the psychosocial factors influencing vaccine hesitancy, in 2011, the WHO Strategic Advisory Group of Experts proposed the “3C” model [4], which stands for “Confidence”, “Convenience”, and “Complacency.” German researchers subsequently proposed the “5C” model, substituting “Constraints” for “Convenience” and adding “Calculation” and “Collective responsibility” [25]. Company-arranged vaccination opportunities are thought to increase people’s confidence in a vaccine, and the availability of the vaccine at or near their workplace increases its convenience. In addition, social environmental factors, such as social norms and herding effects, have been suggested to affect one’s vaccine intention [26,27]. The workplace vaccination program facilitated employees’ vaccination behavior to be shared among coworkers and supervisors, which may have had a direct impact on the social norms and herding effect. Previous studies on seasonal influenza vaccination in the U.S. have reported that workplace vaccination practices and recommendations are associated with higher vaccination coverage [28].

The influence of socioeconomic factors and company-arranged vaccination opportunities on vaccination coverage has implications for the equitable allocation of vaccines. In the workplace vaccination program, the government invited companies that wished to implement the program on the premise that at least 1,000 people could be vaccinated at a single location [13]. Multiple small companies could apply if they could jointly secure more than 1,000 people willing to be vaccinated. However, because it was necessary to arrange venues and medical personnel for the vaccination event and to coordinate costs, program utilization may vary greatly depending on company size. In Japan, employers with less than 50 full-time staff are not obligated to appoint an occupational health physician or health supervisor or to establish a health committee. This lack of obligation means that workers in small companies often do not have occupational health services available to them [17]. In addition, during the COVID-19 pandemic in Japan, there were marked differences among companies of different sizes in the implementation of remote work and infection control measures [14,15]. The Japanese government’s workplace vaccination program may have contributed to health disparities. Therefore, the pros and cons of a workplace vaccination program and the methods used to realize it warrant further discussion to ensure a more equitable implementation in future infectious disease outbreaks.

It is unclear why there was significantly higher vaccination completion among participants who worked for medium-sized companies compared with those who worked for large companies after October 2021 in the model adjusted for both socioeconomic factors and company-arranged vaccination opportunity. One possible explanation is that many employees of large companies were located in offices other than the headquarters and therefore had difficulty accessing the company-arranged vaccination opportunity. Another possibility is that, although COVID-19 vaccination was voluntary, medium-sized companies are often in a weaker business position than larger companies, and therefore they may have been more influenced by pressure from clients to vaccinate their employees in order to continue doing business.

This study had several limitations. First, the survey was conducted via the internet, so generalizations should be made with caution. However, we attempted to reduce any bias by using cluster sampling with stratification by sex, region, and job type. Second, the study was likely affected by recall bias. The earlier vaccination was completed, the more time had elapsed by the time of the survey, which may have caused recall bias. Third, the timing of the follow-up survey might have affected the responses to the question of vaccination status in the last month, December. If a person received their second vaccine dose in the last week of December (after filling out the follow-up survey), they may have answered “unvaccinated” when asked about their vaccination status in the follow-up survey. However, the impact of this situation was likely small because second-dose vaccination was nearly complete in both the community and workplace programs by the end of November, and less than 1% of the respondents received their second vaccine dose in December.

## Conclusion

During the period when COVID-19 vaccinations were being administered to the general population in Japan, the coverage of receiving a second COVID-19 vaccine dose was significantly lower for those who worked for small companies than for those who worked for large companies. This difference could mostly be explained by socioeconomic factors and the availability of a vaccination opportunity arranged by the employer. In the event of future infectious disease outbreaks, it will be necessary to enhance support of vaccination for the employees of small companies.

## Data Availability

All data produced in the present study are available upon reasonable request to the authors

## List of Abbreviations

COVID-19: coronavirus diseases 2019
mRNA: messenger RNA
WHO: World Health Organization
JPN: Japanese yen
OR: odds ratio
CI: confidence interval.

## Declarations

### Ethical approval and consent to participate

This study was approved by the ethics committee of the University of Occupational and Environmental Health, Japan (reference nos. R2-079 and R3-006). Informed consent was obtained from all participants via the survey website.

### Consent for publication

Not applicable.

### Availability of data and material

Data available on request from authors.

### Authors’ contributions

KM designed the analysis, analyzed the data, and wrote the manuscript. TM designed the analysis and reviewed the manuscript. TM created the questionnaire, advised on the study design and data interpretation, and reviewed the manuscript. HA, AH, ST, MT, and SM reviewed the manuscript and advised on the data interpretation. YF was chairperson of the study group, created the questionnaire, advised on the data interpretation, and reviewed the manuscript.

## Acknowledgments

The current members of the CORoNa Work Study, in alphabetical order, are as follows: Dr. Hajime Ando, Dr. Hisashi Eguchi, Dr. Yoshihisa Fujino (present chairperson of the study group), Dr. Arisa Harada, Dr. Ayako Hino, Dr. Kazunori Ikegami, Dr. Tomohiro Ishimaru, Dr. Kyoko Kitagawa, Ms. Ning Liu, Dr. Kosuke Mafune, Dr. Shinya Matsuda, Dr. Ryutaro Matsugaki, Dr. Koji Mori, Dr. Keiji Muramatsu, Dr. Masako Nagata, Dr. Tomohisa Nagata, Dr. Akira Ogami, Dr. Makoto Okawara, Dr. Rie Tanaka, Dr. Seiichiro Tateishi, Dr. Shinya Matsuda, Dr. Tomohiro Ishimaru, and Dr. Tomohisa Nagata. All members are affiliated with the University of Occupational and Environmental Health, Japan. We also thank Katherine Thieltges from Edanz (https://jp.edanz.com/ac) for editing a draft of this manuscript.

